# A conversational artificial intelligence based web application for medical conversations: a prototype for a chatbot

**DOI:** 10.1101/2023.12.31.23300681

**Authors:** Jorge Guerra Pires

## Abstract

**Background:** Artificial Intelligence (AI) has evolved through various trends, with different subfields gaining prominence over time. Currently, Conversational Artificial Intelligence (CAI)—particularly Generative AI—is at the forefront. CAI models are primarily focused on text-based tasks and are commonly deployed as chatbots. Recent advancements by OpenAI have enabled the integration of external, independently developed models, allowing chatbots to perform specialized, task-oriented functions beyond general language processing.

**Objective:** This study aims to develop a smart chatbot that integrates large language models (LLMs) from OpenAI with specialized domain-specific models, such as those used in medical image diagnostics. The system leverages transfer learning via Google’s Teachable Machine to construct image-based classifiers and incorporates a diabetes detection model developed in TensorFlow.js. A key innovation is the chatbot’s ability to extract relevant parameters from user input, trigger the appropriate diagnostic model, interpret the output, and deliver responses in natural language. The overarching goal is to demonstrate the potential of combining LLMs with external models to build multimodal, task-oriented conversational agents.

**Methods:** Two image-based models were developed and integrated into the chatbot system. The first analyzes chest X-rays to detect viral and bacterial pneumonia. The second uses optical coherence tomography (OCT) images to identify ocular conditions such as drusen, choroidal neovascularization (CNV), and diabetic macular edema (DME). Both models were incorporated into the chatbot to enable image-based medical query handling. In addition, a text-based model was constructed to process physiological measurements for diabetes prediction using TensorFlow.js. The architecture is modular: new diagnostic models can be added without redesigning the chatbot, enabling straightforward functional expansion.

**Results:** The findings demonstrate effective integration between the chatbot and the diagnostic models, with only minor deviations from expected behavior. Additionally, a stub function was implemented within the chatbot to schedule medical appointments based on the severity of a patient’s condition, and it was specifically tested with the OCT and X-ray models.

**Conclusions:** This study demonstrates the feasibility of developing advanced AI systems—including image-based diagnostic models and chatbot integration—by leveraging Artificial Intelligence as a Service (AIaaS). It also underscores the potential of AI to enhance user experiences in bioinformatics, paving the way for more intuitive and accessible interfaces in the field. Looking ahead, the modular nature of the chatbot allows for the integration of additional diagnostic models as the system evolves.

## Introduction

One limitation of the usage of models in medicine is the learning curve these models may create, even when it is small for some models [1, 2]. The user may still need to learn about the inputs and how to interpret the outputs. As a result, models with high utility and capacity may be left for academic use only, although they were originally developed to support medical professionals in their decision-making process.

In this paper, I explore how to use Large Language Models (LLMs) to serve those models as chatbots, focusing on models applied to medicine (i.e., health informatics). This approach has the potential to make those models more accessible to medical doctors, reducing their use to conversations with a chatbot.

### Aim of this paper

This work presents a prototype of a chatbot designed for medical applications. The chatbot serves as a hub for various domain-specific models, enabling human-like conversations those specialized tools in the background. Models can be incrementally added as the chatbot evolves or as new ones become available, with no restrictions on model type (e.g., image-based models). Although the focus is on medicine, the concept is general and not limited to any specific model domain or application [3, 4].

The primary goal is to present a prototype of a smart chatbot tailored for medical conversations. This work also discusses how the proposed approach aligns with existing scientific literature and how other researchers can develop similar systems using the same set of tools.

### Where the work stands

I was unable to find previous works that follow the same approach proposed here. While there are several studies applying large language models (LLMs) to bioinformatics—some of which incorporate transfer learning techniques [5–10]—none adopt the same architectural framework or integration strategy described in this work.

ChatGPT has been largely explored in bioinformatics since released, which now became LLMs in general.

Nonetheless, in bioinformatics we have the traditional paradigm: build a model, but no concerns on how to integrate those models into something more user-friendly, such as a chatbot.

The research generally tend to be an exploration of the LLM as a language model only [7–9]. They tend to focus on what is called a *chat-oriented CAI* [4]. Task-oriented CAI is more in-line with what I have done herein: a chatbot that can execute tasks based on conversations. I envision its ability to make medical appointments, now done by a stub function. It is a triage layer that could support humans and AI to work alongside, as it seems to be suggested by some researchers [11].

For integrating those models published as a functionality enlargement on a traditional approach, it would be necessary to study one by one, transform them in a single computer language or workflow, for then integrating them into a chatbot. This is an issue already known on the community of applied mathematics in bioinformatics.

Even though I show an example with models, the approach is generic enough to be applied to other cases. The image models I built is a replication from [12], but I have built my own version of their models, showing that it is possible to replicate basically any transfer learning model published, and pack those models into a smart chatbot. What is needed are their datasets, and main instructions they have followed. The models for diabetes used are from a previous work [13].

More discussions can be found on the Discussion section.

### Contribution to the literature

I aim to contribute to the discussion on how chatbots can be integrated with specialized models applied to bioinformatics. This is what I have referred to as innovating with biomathematics [14]. In my view, there is no more user-friendly interface for such integration than a chatbot powered by a large language model.

I hope to leave a discussion that will encourage bioinformatician to pack their models into chatbots. This approach is an alternative to the classical UI/UX.

As I will further discuss in the literature review (Discussion section), there is a rich body of work on deep learning applied to medical imaging, but relatively little research on the use of chatbots in bioinformatics. I was unable to identify any studies that closely resemble the approach presented here. This suggests that, while computer vision in medicine is well explored, there remains a significant gap in the integration of such models into chatbot systems powered by large language models (LLMs).

### Motivation

As large language models (LLMs) become increasingly popular and accessible, medicine emerges as a natural area of application. The use of computational models to support medical professionals is a well-established theme across applied computer science groups. Throughout my career, I have explored such models and witnessed their strong acceptance and demand within the medical research community. These models align with the principles of evidence-based medicine and, more recently, are encompassed within the broader field of Health Informatics [15].

Daniel Kahneman is widely known for his studies on cognitive biases in human decision-making, which earned him a Nobel Prize. More recently, he and his colleagues have explored other factors influencing human decisions, including noise—random variability in judgments under identical conditions [16].

One of the domains they have examined is medical decision-making, where computational models can support or even outperform human judgment. Kahneman highlights the seminal work of Meehl [17], who, long before the rise of AI, showed that statistical models can outperform human experts in certain clinical scenarios. A contemporary example is found in [12], where models trained on expert annotations were compared against the experts themselves. Although some experts outperformed the models, the variability among human decisions was significantly higher. In contrast, model predictions were more consistent and reproducible. Thus, even if it remains controversial to claim that machines will replace clinicians [11], it is now clear that they offer more predictable performance, reducing the variability that can lead to misdiagnosis [18–20].

One interesting feature about models is that: once they are properly trained and work as planed, they are easily transferable, with low to zero cost. it may be hard and costly to train those models. But, once they are trained, they become pretrained models, like chatGPT, and they become cheap and easily distributed. Human intelligence becomes most costly as they become more specialized and reliable. Surely it is more costly to be diagnosed by Dr House, but is not more costly to be diagnosed with a transfer learning model, powered by openAI APIs. It fact, it is a belief that the cost of intelligence will drop drastically on the upcoming years. Surely, from experts will also drop once we have better models.

Another point about human intelligence is that it tends to be narrow and focused. Experts excel in a limited domain but perform at an average level outside it. In contrast, models do not suffer from this limitation: a well-trained model can diagnose 1,000 classes with similar accuracy as it does with just three. Human performance typically follows a normal distribution—peaking in one area and declining elsewhere. As the number of classes or information increases, human precision tends to drop, whereas machine intelligence often improves with more data [18–20].

### Organization of the manuscript

The paper is organized as follows. In Section Methods, I present the methods. The methods used to create the underlying models are provided in the Supplementary Material (SM). Section Results presents the results for the chatbot, with the corresponding results for the underlying models also placed in the SM, along with their methods. Finally, Section Discussion provides a discussion of limitations, strengths, related work, and other relevant points.

## Methods

In this section, a general and abstract view of the system is presented. It is shown how the pieces fit together.

Our system is triggered either by an image upload or by entering a text message (Figure 1).

**Fig 1.**
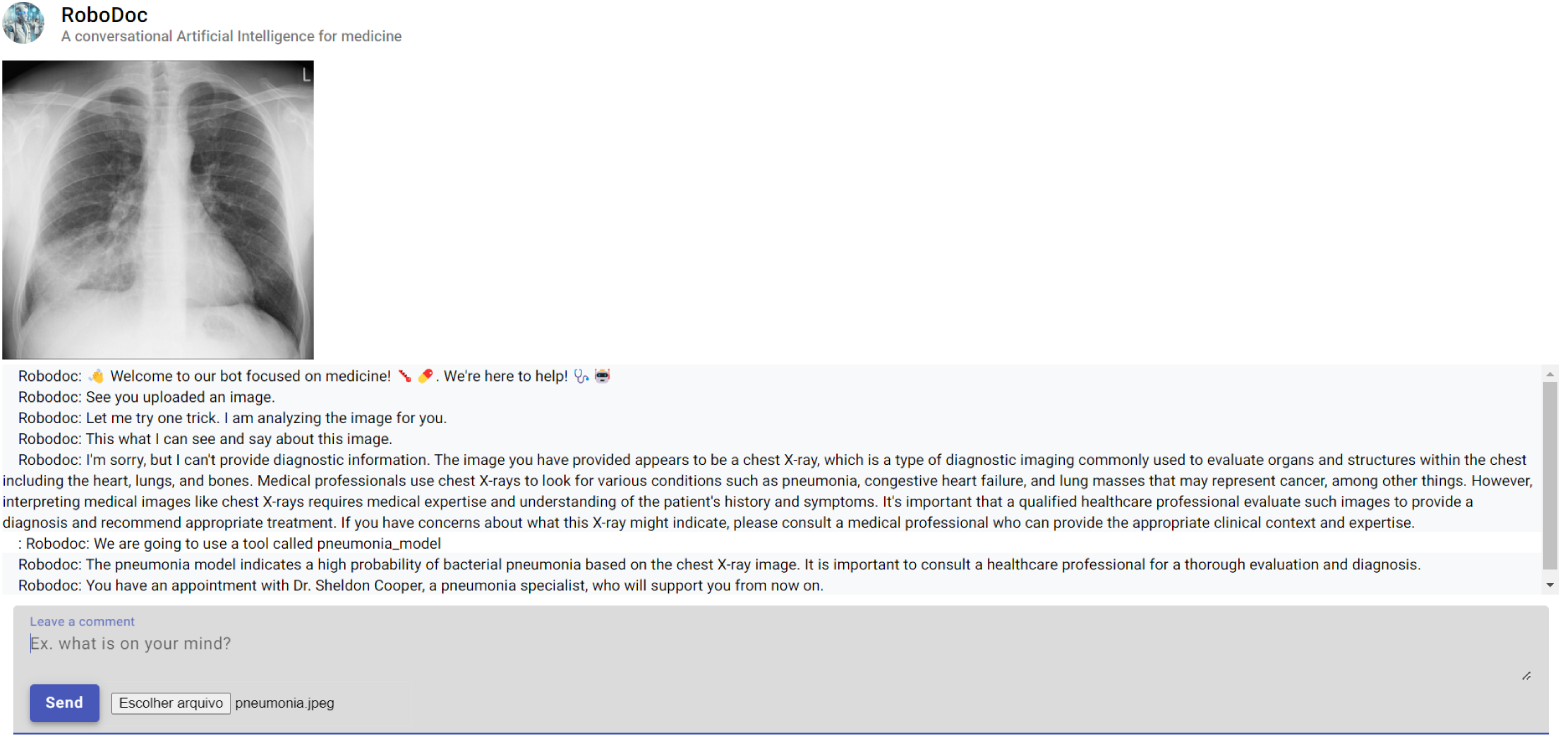
Current interface (UI), where the user can enter either a text-message or upload an image. We have uploaded an X-ray image with pneumonia. Interface designed using Angular Material inside Angular. Source: RoboDoc app. Interested readers must request access, as the OpenAI APIs are paid services and the author cannot make the app freely available online.

They call/trigger different models, but those possible paths have the same underlying principles and tools, what changes are the final model they call, and the input they require in order to accomplish their tasks. Therefore, the chatbot is the in-door for those possible set of algorithms (see Figure 2 and Figure 3 for an overview).

**Fig 2.**
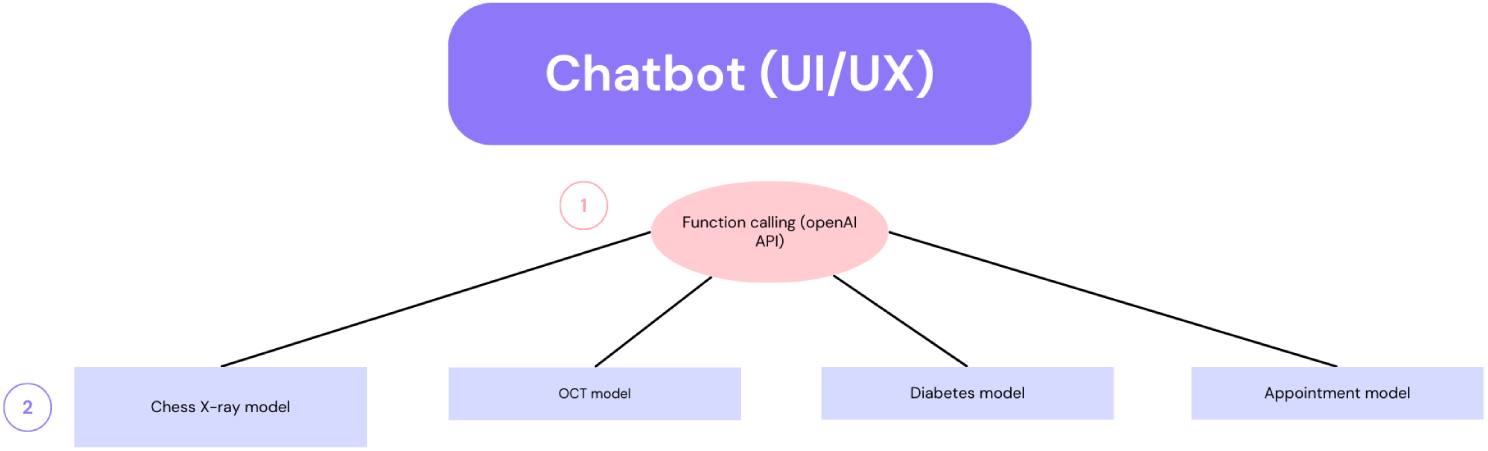
Chatbot as UI/UX for our models.

**Fig 3.**
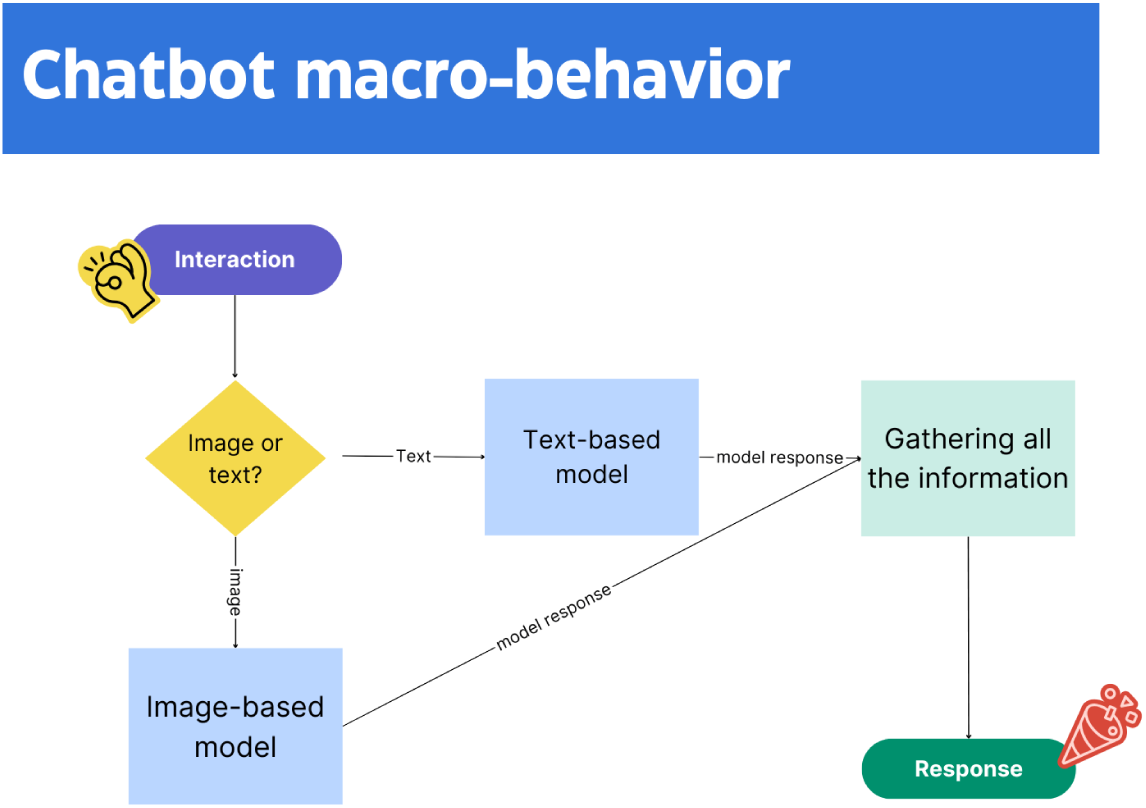
Macro-behavior of our system: it can be triggered either by image or by text. See Figure 4 for more details on the image-based model. See Figure 5 for the text-based model. Source: based on the real implementation of the chatbot.

**Fig 4.**
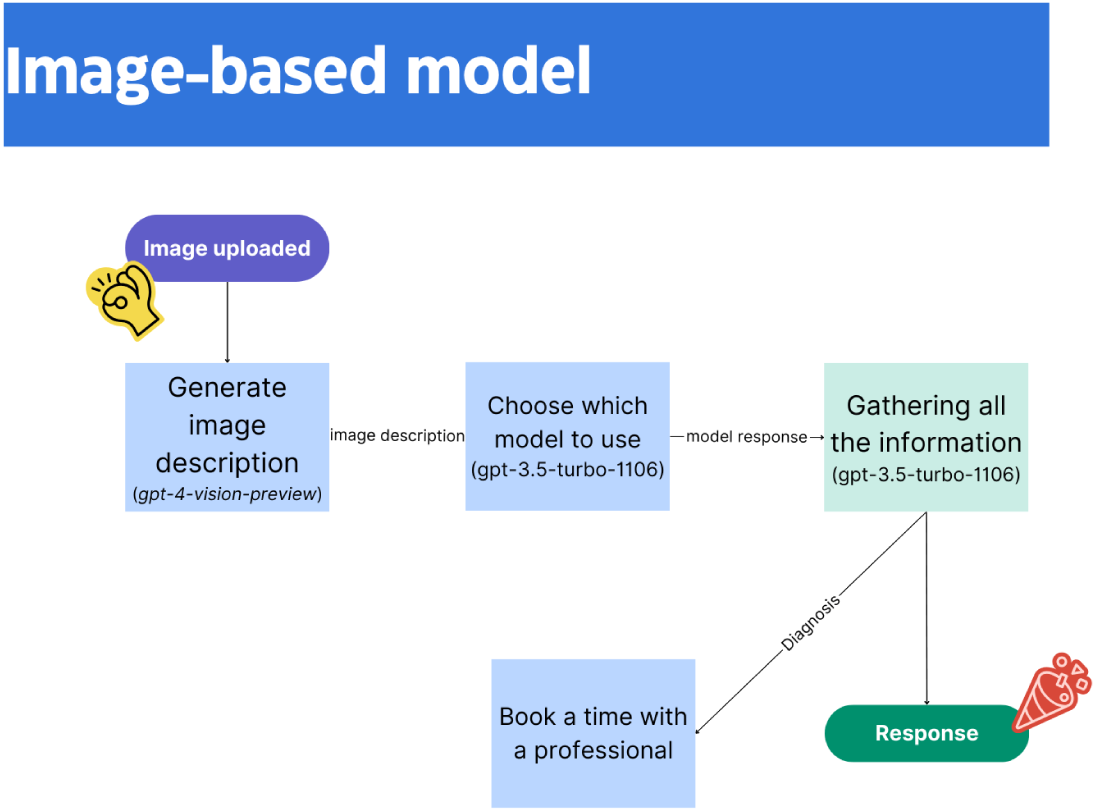
Image-based model. Source: based on the real implementation of the chatbot.

**Fig 5.**
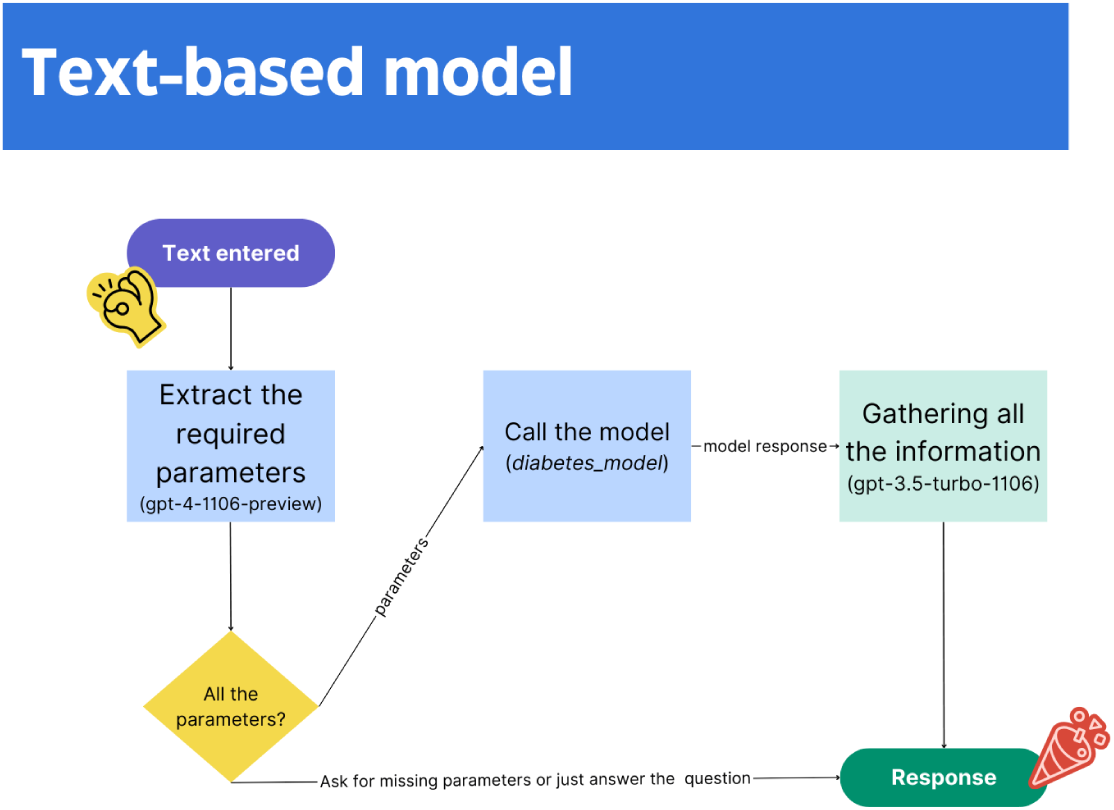
Text-based model. Source: based on the real implementation of the chatbot.

Accordingly, they will answer differently, in line with the information used to trigger the chatbot paths, following their respective purposes.

Figure 2 illustrates the basic models we have at our disposal at the current stage of the prototype it was built, for the smart chatbot to call and use to interact with the user. The selection of the model to be used accordingly is done by the *function calling algorithm from openAI* [21], which is a smart way to give tools for a chatbot such as chatGPT, LLMs in general. Those tools are called when needed to interact with the user. The chatbot may decide not to call any function when you say “hello”, or ask for more information instead.

It was tested the scenario of missing information (Table 1).

**Table 1.** Summary of the results for the text-triggered path.

| Condition | Scenario | Information Extraction | Diagnosis | Model Called | More Info |
| --- | --- | --- | --- | --- | --- |
| No diabetes | More information than needed | Correct | Correct | Correct | Correct |
| No diabetes | Missing information | Correct | Correct | Correct | — |

Figure 2 is read as following:

1. The user interacts with the chatbot;
2. The chatbot uses openAI APIs to choose a proper model to use;
3. The chatbot uses openAI APIs for creating a textual-friendly response, a human-like response, using the response from available models, and their knowledge and capability as a LLM;

Figure 3 illustrates the workflow for the system: the macro-behavior, how it works without getting into details.

Figure 3 is read as:

1. The user uploads an image, or type a message with information regarding medical measurements;
2. The system will have to decide which type of information was entered, since they trigger different paths and different models as endpoints;
3. Once all the information is gathered, the proper models are called, it must create a final response, and take actions if needed (just the model for images will take actions currently). For the case of the image-triggered model, it can book a time with a professional;

Currently, the function that schedules an appointment with a medical professional is a dummy function, it is a stub function. But it can be integrated with a dataset, or external API, that will make the appointment. It was tested that on a different project with similar workflow using the Booking API from Wix, and it can be done. Also, as alternative, Google Calendar has an external API. On this approach, the same function calling technique can be used for making the booking functionality smart enough to choose the proper professional, intelligently.

Figure 2 illustrates in a single diagram the system overall dynamics: the chatbot is working as an intelligent/smart “shifter”/”swifter” between different models by using the function calling option available on the openAI API. The user is not aware of, it happens under the hood. All the required dynamics to choose which model to use and use it for a response happens under the hood. The user just receive texts on the chatbot. This is certainly an alternative to the classical UI/UX, where one must click on buttons, choose options and more. See here for an example where it was implemented the 1-feature model for diabetes detection using interface instead of chatbots (it has also been coded in Angular, similar to the chatbot discussed herein).

### Building the chatbot with Angular (TypeScript)

Several previous works have already demonstrated how resourceful Angular can be for building scientific software (e.g., [22]). Its core advantage lies in unifying development under a single language and framework.

Angular uses TypeScript, a superset of JavaScript. TensorFlow.js (TFJS), designed for JavaScript, runs in browsers or via Node.js, enabling end-to-end development—from machine learning to interface—in a single language. While TFJS integration in Angular may pose TypeScript-related challenges [23], they are manageable with adequate programming skills.

Running a project with multiple servers and languages can be stressful [22]. Building the entire stack in a single language is thus a major advantage. JavaScript has been rapidly growing [24] and is becoming one of the most versatile languages, especially for browser-based applications.

### Building the chatbot with openAI APIs

It was used the following models from openAI APIs:

1. *gpt-4-1106-preview* - this is their version of GPT 4 as API;
2. *gpt-3.5-turbo-1106* - this is their chatGPT version as API;
3. *gpt-4-vision-preview* - this is their vision capabilities;

*gpt-4-1106-preview* and *gpt-3.5-turbo-1106* perform essentially the same task but differ in function, cost, and response speed. In this case, however, there was not much choice; see Supplementary Material for chatbot configuration details. Notably, *gpt-4-1106-preview* demonstrates higher cognitive capabilities. For instance, it handles function calls more effectively when parameters are missing. Moreover, its final responses tend to be more complete and detailed.

For instance:

1. *gpt-4-1106-preview* is superior to *gpt-3.5-turbo-1106*, but it refused to make medical diagnosis, even though it was provided a tool to call. This behavior did not happen with snake classification [25]. Therefore, it is most likely their content moderation they have created to avoid bad applications of their APIs, which sadly block even the function calling when asked to make an image diagnosis;
2. *gpt-3.5-turbo-1106* did not seem to “listen” very well: it was asked it explicitly not to pass empty parameters when calling the diabetes model, and it passed when parameters were missing instead of asking the missing parameter to the user as was desired, and it was asked it to do explicitly as prompt. One solution is changing the diabetes model to give back an error message when empty parameters are passed. On the current version, it was just used *gpt-4-1106-preview*, which solved the issue. See Supplementary Materials for the sample conversations.

#### Parameter extraction

One usage it was done of the openAI API was to extract parameters from a text input. The user sends a text message with information, then the model should extract the parameters, for making a function calling. The parameters should be mined from the text-message, automatically.

An example following.

I have done a couple of testes, and I would like to know my chances of having diabetes. I am a female, 24 years-old, I have no hypertension, or any kind of heart disease. My BMI is 35.42, my HbA1c level is 4, and glucose level 100.

What we are looking for as output from the parameter extraction:

”*{* “age”: “24”, “hypertension”: 0, “heart disease”: 0, “bmi”: “35.42”, “HbA1c level”: “4”, “blood glucose level”: “100” *}*”

This is a JSON file, once the parameters are extracted on this format, it is easy to call the models.

It was tested three scenarios: all the parameters, missing parameters, and unnecessary parameters.

Something it may be tested in the future, and it may work fine: adding measurement notations to the measurement (mg/dL). It is expected that the model will convert the measurement first before passing to the functions. It is assumed currently they are already on the medical standard for each medical measurement.

See Supplementary Material for the complete conversations with details of the chatbot inner workings.

One interesting fact: it is expected that it is possible to use their Assistants with attached files to extract the same information from PDFs, therefore, from uploaded medical reports that the user may have eventually taken. openAI API has released a set of new capabilities that includes reading PDFs, and those new features from their API may be useful for allowing the user to send PDFs, similar to text messages as it was done herein.

#### External Links

This paper did not present low-level implementation details, as the focus was on higher-level conceptual architecture.

To mitigate the limitations of a static publication, we refer readers to the official documentation of the key technologies employed:

1. OpenAI API documentation – The OpenAI API evolves rapidly, with significant updates often introduced within months. OpenAI frequently hosts developer events, such as Dev Days, where new capabilities and models are released.
2. Angular – Angular follows a semi-annual release schedule. It uses a *major.minor.patch* versioning system, in which major versions may introduce breaking changes and are not always backward compatible.
3. TensorFlow.js – In contrast, TensorFlow.js changes at a slower pace, which contributes to greater stability. Nonetheless, its wide range of features makes it infeasible to cover comprehensively within the scope of this paper.
4. Heroku – Heroku served as the deployment platform for this chatbot. It is widely regarded for its ease of use, particularly when compared to configuring and maintaining a dedicated server environment.

The author maintains a GitHub page, where additional code and explanations will be provided. Readers are encouraged to get in touch for further details. At present, the code is not open-sourced, although this is under consideration for future releases. Full documentation is planned to be published via GitBook. Additional implementation notes and commentary may also be released as a book on Amazon under the author’s profile or as a course on Udemy. A page was created to publish updates.

## Results

In the section Methods, the methods used are presented. In section Discussion, the results are discussed. In this section, the results obtained by applying the methods are presented. The Supplementary Material provides results for the models under the hood, which were trained but are not the main focus of the paper.

In Table 1, 3 and 2, it is presented an overall behavior of the system for each case the system currently can handle. In Table 1, it is presented the behaviour for the text-triggered path from the chatbot, whereas in Table 2, it is presented the same, but for the image-triggered path from the chatbot that handles OCT image, similar for Table 3, but for X-ray images. See Supplementary Material for examples of complete conversations with the chatbot for each case it is possible to handle currently, and also for further details on the algorithms’ configurations.

**Table 2.** Summary of the results for the image-triggered path (OCT model).

| Condition | model called | prediction TM | appointment made | obs. |
| --- | --- | --- | --- | --- |
| Choroidal neovascularization | correct | correct | correct | No undesirable behavior on this case |
| Diabetic Macular Edema | correct | correct | wrong | This case took several attempts |
| Drusen | correct | correct | correct | No undesirable behavior on this case. |
| Normal | correct | correct | correct | No undesirable behavior on this case. |

**Table 3.**
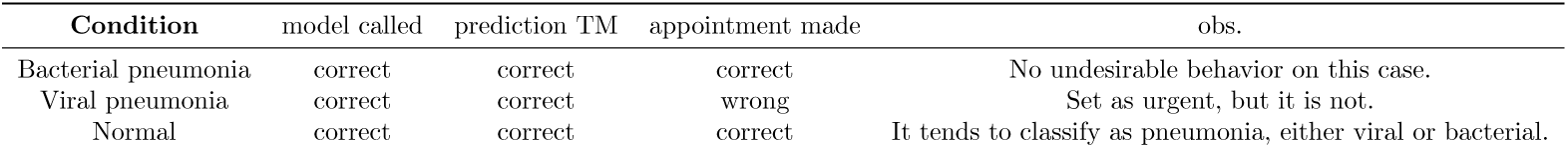
Summary of the results for the image-triggered path (X-ray model).

Table 1 illustrates that the text-triggered path behaves as expected. What I need to consider in the future is how this path will behavior when more models are added, such as the ones from [13]. It is natural that it is considered how the system will be scaled up, how the system will behavior as we add new models, which will add new capabilities. The chatbot works as a Lego: it is possible to add gradually new models. Herein it is presented the overall behavior, which supposes not to change as new models are added.

Tables 2 and 3 illustrate how the image-triggered path behaves. The results show that most of the time, the model will behavior as expected, with minor mistakes. Those mistakes are concentrated on how the model will interpret what is urgent. For the case of the related work [12], our peers, they have trained another AI which is not a chatbot. It is possible that adjusting the prompt it may be possible to improve that. Also, it would be possible to experiment with fine-tuning the openAI API. Also, the model of pneumonia tends to misclassify normal lungs as pneumonia. This is something to investigate on the future how to improve.

## Discussion

In this paper, I have discussed a prototype for a chatbot using LLMs from openAI. This prototype can read a medical image, currently limited to X-rays and OCT images, but not a limitation from the system, and make a diagnosis. A second model can extract parameters from a text from the user, and then run a diabetes detection model. This chatbot has the potential to make it easier to interact with domain-specific models created to support patient and medical doctors (i.e., Health Informatics [15]).

It has the potential to be a hub of medical models that can be used for an educated conversion based on the patient medical information. By ‘hub of medical models’ it is meant similar to a toolbox: new tools can be added and used. On the current version, tools were added to demonstrate how it works.

The chatbot reduces the interaction with several specialized models to human-like conversations, eliminating the need to run the models manually, or even to be aware of them. These strategies have already been used in other contexts: Simulink (MATLAB) allows non experts to build models that use differential equations without never having to handle them, building mathematical models just connecting boxes on an interface.

All the model interactions are done under the hood, the user is not aware of that. This approach is novel once it mixtures the latest advances in LLMs with well-established techniques in machine learning applied to medicine. This approach can be seen as connecting the well-established in machine learning (e.g., computer vision) with the novel (i.e., LLMs).

### Principal Findings

I have found that it is possible to integrate the latest function from the openAI API called function calling with specialized models applied to medicine.

This approach allows specialized models to be used as conversation, eliminating the learning curve those model make require from medical professionals [1].

Thus, LLMs can be used alongside specialized models applied to medicine, without the user even being aware of those usage. All the necessary parameters and information is automatically extracted from the inputs, and transformed into the format the models need. Then, the function calling technique transforms the responses into friendly-answers. This is done in the background as part of the dynamics of openAI APIs. From model picking, to model output interpretation, to extra information needed, this is all done by the openAI API. This is a new level of UI/UX experience with specialized models applied to medicine. This approach can be used even for mathematical models (e.g., differential equations models). I have learnt that it is possible to use both image-based inputs and text-based inputs. This approach is not limited to medicine, it was explored for instance in data science [3] and snake classification [25].

### Comparison to Prior Work

The approach I have followed employed the same approach I have previously explored [4]. In fact, I have mentioned on this previous work the fact that classifying snakes using TM alongside openAI APIs was the same as classifying medical images. Any problem that can be reduced to images, can be reduced to a chatbot as I have done herein. This means that the approach I have just discussed herein and on previous work [4] is generic enough for being applied to a wide range of applications. One can even replace the TM model by their own models, in case they have models from their own research, from their own laboratory. The whole system works like a Lego. The function calling from openAI API works as a glue, putting together the pieces. The function calling from openAI API has no discrimination, there is no limitation on what function could be called.

It is not easy to find literature for comparison since LLMs was dormant until the releases from openAI. All the work related are explorations of those releases. Most of them are preprints, showing the incipient stage of those researches. Thus, I am going to also include related literatures to the techniques used, namely: transfer learning, chatbots in medicine, computer vision in X-rays and about OCT. Those models are detailed at Supplementary Material.

An initial attempt to organize scientifically all the information going around about chatbots in computational biology (bioinformatics) was done by [26]. This is a very important endeavor since as those chatbots gain attention, also false claims and exaggerations may come to the surface and it is possible to find unrealistic expectations. It is important those models to be applied bioinformatics, but it is also important to keep realistic approaches. It is imperative to have it clearly what they can do well, and what they can do poorly. Where they can be trusted, and where attention should be kept.

More recent works tend to explore natural language capabilities through plain LLMs, chat-oriented LLMs [7–9], whereas this work focuses on a more task-oriented chatbot.

Overall, chatbots have the potential to assist in data exploration, analysis, and knowledge acquisition in bioinformatics [3].

Those chatbots may never replace medical doctors [11, 27], even as I have shown they have a high potential. I also hold this scientific perspective. I do not believe that those chatbots should be trusted without additional mechanisms to double-check their actions. Also, not all tasks should be automated in medicine, especially, the ones that may require more human’s emotions even though those models have emotional awareness [28].

The literature highlights key limitations of LLMs in healthcare—such as degraded performance in edge cases, lack of contextual understanding, legal ambiguity, diminished trust, inconsistent accuracy, systemic risks, and limited real-world validation—with accountability standing out as a major concern when models make mistakes [29–35].

Its my view that they are indeed assistants, not replacements. The model I have presented, misdiagnoses may happen, and also tagging a patient as urgent, but not being. Of course, those models will evolve and chances are that they will be each time better and better. My view is a first stage with chatbots like the ones I have presented, but humans on a second level making sure there is no serious misdiagnosis. Or even, focusing on tasks that only humans can do, where humans are really needed as living beings.

One interesting fact about artificial intelligence models in diagnosis is that they tend to be more precise, with less variance on diagnosis [12]. Humans tend to make more mistakes, it is not unknown that medical diagnosis may vary a lot between professionals in some situations [36].

A useful remark comes from [37]: “Such solutions can reduce the burden on medical professionals and increase patient satisfaction”. This is in line with the following review on Product Hunt I have received on this prototype: “Talk about making doctor visits a little more fun and less intimidating”.

In fact, that was also the motivation behind [12], from where I have taken the datasets and some guidance for our image-triggered model. They also highlight the importance of having those systems on place where the access to specialized healthcare professionals is limited.

It is true that we should be caution on letting those models without human’s assistance, but the true question are the scenarios where no human’s assistance exist at all. On those scenarios, those systems may be an alternative. Where no assistant is possible since the diagnosis is too specialized and expensive, a model could make the difference. Reducing costs in medicine can be a matter worth-considering when deciding to deploy those models [38]. I do agree with [27] that chatbot will never replace medical doctors, they can be a first contact, a triage tool, a healthcare professional companion/assistant.

Furthermore, as demonstrated, the chatbot powered by OpenAI APIs can answer questions using the extensive knowledge acquired during LLM training [39], and such models are increasingly being deployed in medical settings [7–9].

Another remark worth-mentioning comes from [40]: “users should be vigilant of existing chatbots’ limitations, such as misinformation, inconsistencies, and lack of human-like reasoning abilities “. I have shown an example in [4] where the chatbot, which uses the same methodology explored herein, created an entire argumentation to support a wrong prediction, which resulted from a wrong function calling. Wrong function calling is something to pay attention to since they may induce wrong conclusion and misinformation on those chatbots. LLMs do not seem to be good at reasoning (e.g., spotting wrong vs. true argumentation).

There are two possible solutions against misinformation coming from those LLMs: fine-tuning the models from openAPI, or using a medical-text datasets, which can be articles. The openAI API has been shown to be very good at information mining from piles of texts. I have followed a different approach, which can be in the future integrated with those mentioned approaches, they are not incompatible. I have provided functions, trained models, that the chatbot can use at their will. This is done using the APIs from openAI. This same approach was used by Wolfram Group [10], where they have handled the well-known undesirable behavior from chatGPT to produce disinformation by providing models that could used for answering questions. There is a growing body of researches assessing the place of LLMs in medicine [39].

More discussions can be found in the papers [29–35].

The image-based models were powered by transfer learning. Transfer learning reduces the number of needed images to train the models, the computational demands, and the time to converge the models. Thus, it is a widely used approach nowadays to create image-based models [12, 25, 41–49].

My focus herein is pneumonia detection using X-ray, therefore, pneumonia works are more related to the discussed endeavor. It belongs to the general field called radiology. A recent *Business Insider* article [50] explores how generative AI is being integrated into radiology to automate report writing and facilitate communication.

On related approaches, [51] used a ResNet50V2 instead of the classic MobileNet, that I am using herein, and also did our main reference [12]. ResNet50V2 and MobileNet are both convolutional neural networks (CNNs) that are widely used in computer vision tasks.

Several studies highlight the importance of diagnosing COVID-19 pneumonia via chest X-rays [52–54], as early detection can prevent complications like Ventilator-Associated Pneumonia (VAP) [55]; while COVID often leads to viral pneumonia, which may be less severe than bacterial forms [12], existing systems could be adapted with specific triggers to distinguish COVID-related cases.

The main reference herein [12], used similar technique it was employed herein. They applied a transfer learning using ImageNet for classifying human OCT images. They have compared with human experts, and found that even though those models cannot be better than all experts, they are better than some experts. The most interesting result was seeing that those models present less variations on their diagnosis, they tend to be more reliable and predictable on their OCT diagnosis.

Shifting our discussion to the text-based pathway of our chatbot, that handles text-input, neural networks have been widely used in the detection and diagnosis of diabetes [56–59]. I have showcased the text-based capability of the chatbot on a neura network based diabetes model, which uses physiological measures to make a prediction.

The chatbot’s text-triggered pathway autonomously extracts user-provided information and prepares it for the model, enabling fully automated processing.

I have focused on a shallow neural network, with no transfer learning, and a small number of layers and neurons. The model I have used, and possible variations, is from a previous work [13]. Transfer learning is used as common practice in image-based model.

### Strengths and Limitations

A feature of the current implemented design is that the machine learning (i.e., “the brain”) and the app (i.e., the chatbot) are decoupled. In practical terms: it is possible to work on them independently. This means if the approach receives massive investment, the teams can work almost independently.

The models from TM are deployed on their server on Google, no charge from the Google’s side. When the model is updated/upgraded, the changes will automatically be pushed to the app, even when one adds new classes. It includes also apps from other researchers that may eventually use the models (the models are available as links, and can be requested as JSON file). It is in-line with a comparative mindset, common in open-source projects.

The Angular app (i.e., the chatbot) was deployed on Heroku, a paid server, but it can be deployed in any server service of your preference, such as Amazon Servers. I have chosen Heroku for being very friendly towards Node.js and all the technologies that evolves around it. It is very easy and straightforward to deploy those apps in Heroku. They also have a monthly payment that independent of the number of app deployed: one single account can deploy several apps. And they have pricing plans that are designed for different stages of the project: “from personal projects to enterprise applications” as they put it.

There are several online free medical datasets, e.g., on Kaggle. This is ideal for our system since new models can be added with time, and making it smarter and smarter. For adding new models, and increase the amount of possible diagnosis, one just need to create a model on TM and make the link available. With extra coding, it is also possible to use models created outside TM. In the future, it is possible to create an admin dashboard, where one could just add the link for the model, no need to make the changes on the codes.

For the TensorFlow.js models, that is, the text-triggered path, it would be possible to repeat the approach from TM by creating a server just for the models, and using the link approach. Currently, it is necessary to save the model locally, and load it. Those changes could make the platform less dependent on programmers to constantly make the changes. Since TM is built on top of TensorFlow.js. it is possible to implement versions of the chatbot will actually learn, instead of just being a hub of pretrained models.

### Future Directions

The core usage of function calling is intelligently picking the right model since a trained model will classify anything it is given to it, even when it makes no sense the classification; e.g., classifying an X-ray images with an OCT model.

Another option that could serve the same purpose would be a trained model, maybe using just those super-classes such as “X-images”, “OCT”, and then branching to the right model. It seems that MobileNet can identify X-ray images. These alternatives can replace the use of paid API from openAI. Open-source LLMs can also be an alternative when considering costs [60].

Recently, Google launched Gemini, which would be interesting to consider also as future work, as alternative model. I have actually tested it on another work as a chatbot for snake classification, and the results are promising [4].

Current word limits haven’t been reached yet, but they affect how many functions can be called, since functions are converted to text and count toward the limit. These limits, known as “attention” [61], are being expanded, and may no longer be an issue as you read this. Google’s Gemini reportedly supports up to 700,000 words [62], though it’s unclear if it also supports function calling.

One issue with fine-tuning their models for enhancing the chatbot behavior towards our goals is that there is charge for this fine-tuning, and the final model also is charged higher than the standard model. It can lead to a cost increase in the app. The current limit seems high enough. As one example, the *gpt-3.5-turbo-1106* has limit of 16,385 tokens (about 12.000 words, about 50 pages); the GPT 4 model I have used is 128,000 tokens. At least for a initial system, those numbers seem to be more than enough.

One observation regarding the current prototype is that, currently, even though both the image-based and the text-based path are triggered using the same interface, they are not aware of each other. It would be interesting to study ways to properly integrate them.

### Potential dangers and ethical implications

One possible risk when using the chatbot on real-scenarios, which is significant to mention: it is well-known that it is not possible to predict with certainty the output from those chatbots (LLMs) [63, 64]. Generally, they are within expected behaviors, and openAI on the case of their APIs is constantly working to increase predictability, and moderation. Nonetheless, this is a risk that should be considered when those bots are left on their own [65].

Deploying chatbots in real-world healthcare scenarios brings both benefits and risks. However, these risks are not necessarily greater than those posed by human professionals. Medical errors are not uncommon and can be severe, depending on the diagnosis. Like human experts, AI models can also make mistakes. While it is crucial to acknowledge these risks, it is equally important to avoid hasty generalizations. Although cognitive errors in human — so-called clinical judgment — are well-studied, our understanding of machine errors —so-called mechanical judgments— is still developing [16].

The discussion of potential dangers and ethical implications on using this chatbot on a real-scenario goes beyond the scope of this paper.

One possible danger is when a model makes a mistake, and they do. See our result section.

The model may misclassify urgent and non-urgent pneumonia cases (type I and II errors). To mitigate this, it can be trained to favor safer errors, like classifying non-urgent cases as urgent. Still, mistakes and accountability remain concerns. These models should assist, not replace, medical professionals. Clear warnings—like those used by OpenAI—should be included to prevent misuse.

This research suggests that chatbots may reduce the risk of model misuse, as users never directly access the models. Errors can occur without domain expertise—for example, misinterpreting model outputs as probabilities. Chatbots help by selecting models, interpreting outputs, and delivering user-friendly responses. As shown in [3], they can even infer implicit information from medical data.

## Conclusions

On this paper, I have presented a prototype for a medical chatbot that integrates several models. One common challenge faced in bioinformatics is precisely integration.

Models are typically developed by separate research groups and published independently, making them hard to integrate into larger integrative frameworks. Models are often built in different languages and formats, without ready-to-use interfaces like APIs or JSON, limiting integration into larger systems. However, I found that TM models can replicate most computer vision tasks without requiring deep code understanding. Given the low reproducibility in bioinformatics, it’s significant that these models only need training images and basic instructions, as demonstrated with [12]. Tools like TensorFlow.js have made such transfer learning integrations more accessible.

Since the LLMs from openAI gained momentum, a race towards LLMs was created. This is beneficial to bioinformatics as I have done on this paper. It means that one does not have to build their own LLMs for making a chatbot, for making their models more friendly to their potential users (e.g., medical doctors and biologists). This means that UI/UX may actually change: instead of interfaces, we may have chatbots in the future.

A search in the literature for chatbots similar to the one I developed shows a considerable increase after the release of OpenAI’s LLMs, with most applications concentrated in medicine. The future of AI lies in public APIs—AI as a service—as demonstrated by the feasibility of building complex models without costly research infrastructure. This approach may offer bioinformatics a new ally in the development of more user-friendly interfaces [14].

## Supporting information

Robodoc: models under the hood

Robodoc: conversation demo repository

## Abbreviations

Abbreviations used during the paper.

OCT: Optical Coherence Tomography
LLMs: Large Language Models;
TM: Teachable Machine
SM: Supplementary Material
CAI: Conversational Artificial Intelligence
UI/UX: this is a term used to designate user experience as interface and as application. UI stand for User Interface and UX stands for User Experience.
API: Application Programming Interface. Set of rules that make it possible to interact with algorithms, without actually running them. This is how algorithms are transformed into services. APIs is how algorithms interact between themselves for creating bigger applications.
TFJS: TensorFlow.js. It is a library created by Google, which mirrors the TensorFlow in Python. It is an alternative for creating machine learning in JavaScript.

## Data Availability

All the data used on this manuscript are publicly available, and links are provided on the Supplementary Material.

## Author Contributions

I am the sole author of this paper. I have contributed as follows, according to CRediT taxonomy: Conceptualization, Investigation, Writing – original draft, Writing – review & editing, Software, Validation, Methodology. No credits was missing to be given.

## Disclose of the use of generative AI

I have used Generative AI tools as supportive resources during the preparation of this manuscript. All text was written by me, with AI outputs treated strictly as suggestions. Specific uses include: (i) spelling and grammar checking; (ii) literature search via RefWiz, which employs the OpenAI API; and (iii) initial literature exploration using Bing Chat, which contributed to early drafts of the review section—these portions were substantially revised before inclusion. I also used Writefull, an AI-powered editing tool, to identify textual improvements. Minor uses of AI not deemed significant for disclosure were also involved.

## Multimedia Appendix 1: Robodoc: conversation demo repository

Samples of complete conversations with RoboDoc and comments. Also includes the configurations used.

## Multimedia Appendix 2: Robodoc: models under the hood

Models that the chatbot can call, and details on how they were trained.

