## Supplementary material for "A conversational artificial intelligence based web application for medical conversations: a prototype for a chatbot": Robodoc: models under the hood

#### Abstract

This document provides details on how I have trained the models used to showcase Robodoc. For the chatbot, please, see the main paper.

**Keywords**: chatbots; bioinformatics; conversational-AI; openAI; large language models; tensorflow.js; JavaScript; medicine; medical images; computer vision.

#### 1 Introduction

The chatbot I have discussed on the main paper is powered by models that the user does not see. On this appendix, it is possible to see how those models were trained, and their behavior. See the main paper for seeing how the pieces fit together to create a smart chatbot.

#### 2 Methods

See the main paper for the methods on the chatbot. On this section, I have separated the methods for the models used to showcase the chatbot, the models under the hood.

##### 2.1 Diabetes model: building models with TensorFlow.js

The model we have (re)-built was discussed on Pires (2023), and also variations of this model. The model files, the model ready to use, can be downloaded from GitHub.

One can upload the model in Angular, or make the proper adjustments for their preferred computer language, using Angular (TypeScript):

```
async diabetes_model(features: number[]){

  //Location of the model, should be downloaded
  //The other file should be placed in the same folder
  const modelPath = './assets/my-model.json';

  let output: any = {message: "no prediction done"};

  //Loading the model locally
  return tf.loadLayersModel(modelPath).then((model) => {

        const input = tf.tensor2d([features]);

        //Making a prediction using the diabetes model,
        //already trained
        const result = model.predict(input) as tf.Tensor;

        //TensorFlow.js stuff,
        //needed to get the values from GPUs
        const prediction_value = result.dataSync()[0];

        output= {probability: prediction_value};

        //This is needed for openAI APIs
        //to understand the response as string
        return JSON.stringify(output);

      })
  }
```

TensorFlow.js was created mirroring TensorFlow in Python; one can even transform models in TensorFlow to TensorFlow.js, and use them normally Laborde (2021), and *vice and versa*. We have trained our model on an extensive dataset on diabetic and non-diabetic patients, available freely on Kaggle. It was used 2.000 samples, equally distributed between diabetic and non-diabetic patients. The model had to predict whether a patient had diabetes based on six features (Table 1).

It was deployed here a version of the 1-feature model from Pires (2023), just the interface for the model, no chatbot. This is the alternative to the chatbot as interface.

##### 2.2 Building the models with Teachable Machine

Teachable Machine (TM) is a platform created and maintained by Google. It is not clear from their official documentation how it works, but it is known they are using TensorFlow.js (TFJS) Laborde (2021), an API for creating neural networks focused on deep learning, also created and maintained by Google. It possible

**Table 1:** Features used to train the model with six features for diabetes detection.

| Feature | Short Description |
|---|---|
| HbA1c level | Higher levels indicate a greater risk of developing diabetes. |
| age | age ranges from 0-80 in our dataset. Diabetes is more commonly diagnosed in older adults. |
| bmi | BMI (Body Mass Index) is a measure of body fat based on weight and height. Higher BMI values are linked to a higher risk of diabetes. |
| blood glucose level | Blood glucose level refers to the amount of glucose in the bloodstream at a given time. High blood glucose levels are a key indicator of diabetes. HbA1c level is a long term measure, 2-3 months. |
| heart disease | Heart disease is another medical condition that is associated with an increased risk of developing diabetes |
| hypertension | Hypertension is a medical condition in which the blood pressure in the arteries is persistently elevated. |

Source: Pires (2023)

they are using additional metaheurics that increases the platform performance, compared to just using TFJS, as it is. Metaheuristics can make the difference on those algorithms. It is possible to learn from the source code, which is open source.

An apparent superiority of TM models compared to our peers were noticed Kermany et al. (2018), but it hard to explain why since the technical details of TM is not clear on their official documentation. It is possible to infer from their official documentation how it works from a general standpoint, but not their inner workings. It is possible that those details are on their GitHub repository. Those details will be left for the reader to explore, just the platform is explored herein.

It is possible that metaheuristic they have created on top of TFJS, which gave their models this apparent superiority. By superiority, we mean: i) the models seems to converge faster than the ones reported in Kermany et al. (2018), theirs take hours to converge, whereas ours take minutes, no more than 1-10 minutes for converging; ii) TM seems to require lesser images (about 30 images for each class, which is a very small number).

By reading the paper of our peers Kermany et al. (2018), it seems the same approach that was applied herein is explored on this paper, which makes is harder to associate any possible superiority on the models. They are using transfer learning using ImageNet as dataset: "Using the Tensorflow we adapted an Inception V3 architecture pretrained on the ImageNet dataset " Kermany et al. (2018). Inception V3 is a more complex architecture that is better suited for high-performance computing environments, while MobileNet is a lightweight architecture that is more efficient for mobile and embedded vision applications. For sure working on the browser, it is better to travel light. Inception V3 was introduced in 2015 by Google researchers, whereas MobileNet was introduced in 2017 by Google researchers.

Another scientific inquiry that we were unable to confirm: ImageNet may have changed since their publication in 2018; maybe also the feature models, since in artificial intelligence, models may become obsolete fast. We have also noticed this patterns on another research we did Pires and Dias Braga (2023). We actually tried to replicate this behavior, using just TFJS, but we could not see the same superiority of the final algorithm in term of convergence.

Those are all possible speculations.

It is straightforward to create a model using TM:

i. Open their platform;
ii. Choose your model configurations, very basic;
iii. Add the classes;
iv. Add the images per class;
v. Ask to train;
vi. See the metrics they provide after training;
vii. Export your model either by downloading or uploading the model to their cloud;

We have chosen to upload the model to their cloud. The model will be available as a link. If this link is used on the browser, you will see an interface with the model; if the link is used on a code, it will upload the model locally. Below is an example on how one can upload locally a model from TFJS, and make a prediction.

```
//Loading the model locally
 const modelURL = TFlink + 'model.json';
 const metadataURL = TFlink + 'metadata.json';
 const model = await tmImage.load(modelURL, metadataURL);
const prediction = await model.predict(image);
```

What is interesting regarding this approach: adding new models is relatively easy. Therefore, one can add new models as soon as they have a new image dataset, and the changes on the core model will be minor. The chatbot will have the chance to call new models as soon as it is available to be called. On this approach, a very complex chatbot for medicine can be built by parts, using a "Lego approach". Nowadays, it is possible to find several online medical datasets for free (e.g., Kaggle list).

At the current moment, TM does not support datasets like the ones for diabetes: it is focused on image, videos and sounds. That is why we had to build the diabetes model using TFJS directly.

TM uses *transfer learning*, the same underlying approach from our peers Kermany et al. (2018). That is why you can train a model with 30 images in TM, a typical image model may require millions of images. For Kermany et al. (2018), even with transfer learning, it took them hours to weeks to train their models. TM takes minutes to finish the training; for 30 images, it takes seconds.

#### 2.3 Datasets

On this section, we discuss the datasets we have used for training our models. All the datasets are public, and available on Kaggle.

##### 2.3.1 *Pneumonia model*

The complete dataset can be found on Kaggle here. The reduced version we have used can be found here on Kaggle.

##### 2.3.2 *Retinal model*

The whole dataset can be found here on Kaggle. The dataset was firstly modeled and made available by Kermany et al. (2018). We shall use this publication to compared our results. For your convenience, we have saved the exact dataset we have used, a reduced version from the original here on Kaggle.

The dataset is divided into four classes, three of them being medical conditions on the retina:

i. Choroidal Neovascularization (CNV) (1245 images);
ii. Diabetic Macular Edema (DME) (888 images);
iii. Drusen (1064 images);
iv. Normal (1092 images);

The image counts are for our dataset, not for the original one. The numbers were decided "randomly": we ran simulations with different numbers, those seemed to give a trade-off between amount of images per class and quality. When using a small number of images (about 30 images per class), the model will converge even better and faster, and when tested with "random" testing images, it seems to generalize. When using many images, it is actually possible to calculate some basic metrics, that is: confusion matrix and accuracy. Having those metrics on many images is statistically more significant than having for a small number of samples assigned for the testing dataset. The testing dataset size is proportional to the number of images per class. Generalization may be more effective if we have more image.

Those are the classes we must spot, learn from the dataset. The AI sytem should give special attention to "images with choroidal neovascularization [CNV] and images with diabetic macular edema [DME] as 'urgent referrals'." Kermany et al. (2018). Thus, we need to make sure those cases are treated with special attention. Those two cases can lead to blindness.

#### 2.4 Computer resources

All the simulations were done in the browser, and they did not last more than five minutes each. The computer main configurations were: Windows 11, Vostro 7620 Dell 12th Gen Intel(R) Core(TM) i7-12700H, 2300 Mhz, 14 core(s), 20 logical processor(s).

### 3 Results

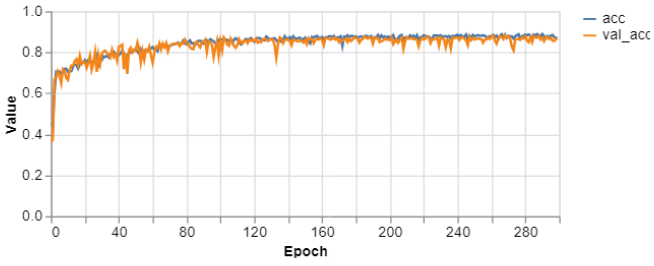


**Figure 1:** Accuracy for the diabetes model. Training in blue, and validation in orange.

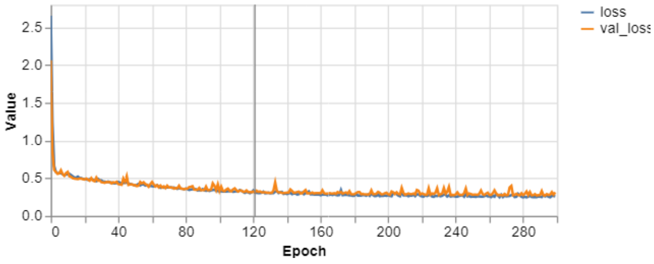


**Figure 2:** Loss function for the diabetes model. Training in blue, and validation in orange.

#### 3.1 Diabetes model

Fig. 1 illustrates the training process, the accuracy as metrics. The training process converged to about 90% for both validation and training curve. It means that the model generalized, and was able to learn the pattern on the dataset provided. This curves suggest that there were no overfitting, or underfitting.

Fig. 2 illustrates that the loss function for both training and testing converged to lower values. Alongside Fig. 1, it shows that the model learnt the relationship between diabetes and the features used.

#### 3.2 Pneumonia model

Fig. 3 illustrates the accuracy per class for the pneumonia model. The highest accuracy is for normal x-ray image (97%), whereas the hardest case is for virus pneumonia (66%).

Fig. 4 illustrates the confusion matrix for the pneumonia model. The highest misclassifications happen between virus and bacteria pneumonia. Kermany et al. (2018) focused on a binary model: pneumonia vs. normal lungs. What is interesting on this matrix is that most of the misclassifications are on the virus-bacteria sub-matrix (upper corner on the left). This means that the model tends to make mistakes amongst pneumonia types. Similar results was found by Kermany et al. (2018), as a consequence, their model is focused on the binary classification: pneumonia vs. normal.

Fig. 5 illustrates the accuracy of the model. Both the validation curve in orange and the training curve in blue converged, about 80%-100%. Therefore, the model was able to learn, and generalize. Those results point out that

Accuracy per class

| CLASS | ACCURACY | # SAMPLES |
|---|---|---|
| Virus | 0.66 | 202 |
| bacteria | 0.80 | 271 |
| Normal | 0.97 | 202 |

**Figure 3:** Pneumonia detection, accuracy per class. Source: own results using Teachable Machine.

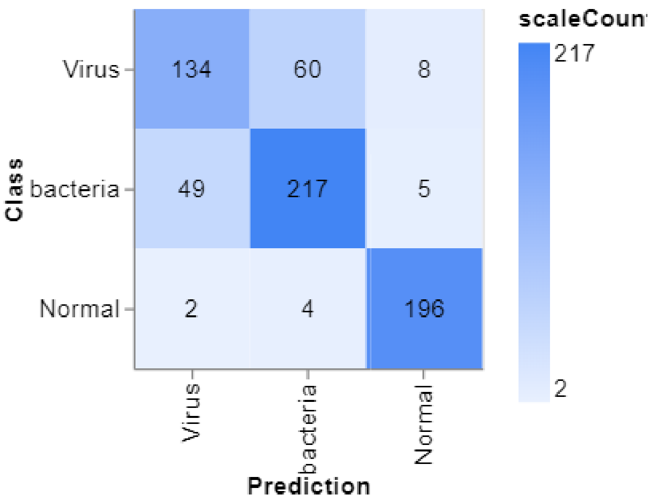


**Figure 4:** Confusion matrix for the pneumonia model. Source: own results using Teachable Machine.

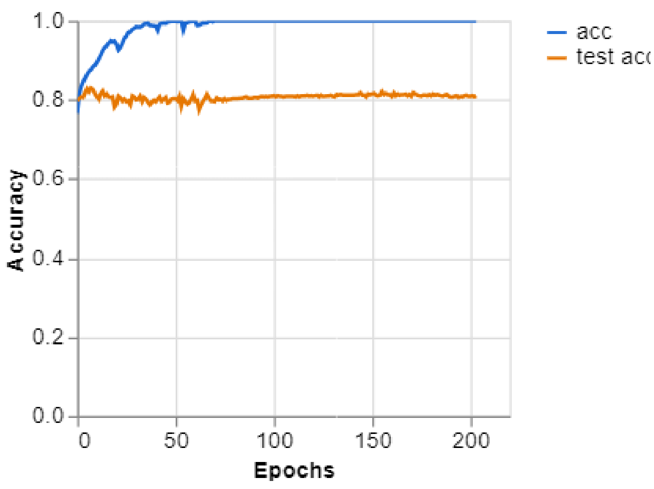


**Figure 5:** Model accuracy during training for pneumonia model. Training in blue and validation in orange. Final: 81% for testing (orange) and 99% for training (in blue). Source: own results using Teachable Machine.

no overfitting or underfitting most likely did not happen.

Fig. 6 illustrates that the loss function for both training in blue and validation in orange converged to low values. This result points out that the model learnt from the dataset presented.

The final model can be found here.

#### 3.3 Retinal model

Fig. 7 illustrates the four possible classes for the OCT images entered, and their respective accuracy per class. The lower accuracy is 88% for Drusen, and higher is 98% for CNV.

Fig. 8 illustrates the confusion matrix. Most of the classification were true positives, whereas some false positives can be found. Similar result was found by my ?.

Fig. 9 illustrates the accuracy of the model. Both the training in blue and validation in orange converged.

Fig. 10 illustrates the loss function for both training in blue and validation in orange. Both curves converged to low values, and remained low.

You can find the model on this link. This model is stored on a Google Cloud, as courtesy from Google, once a model is trained, it is possible to upload it to their cloud as part of their features for Teachable Machine. It is possible to upload an image and test the model using this link directly on the browser. As alternative, it is possible to use the same link to upload the model locally, and run in your application. That is what I am going to do: I am going to load the model on my Angular code, using this link, for making it available to the chatbot.

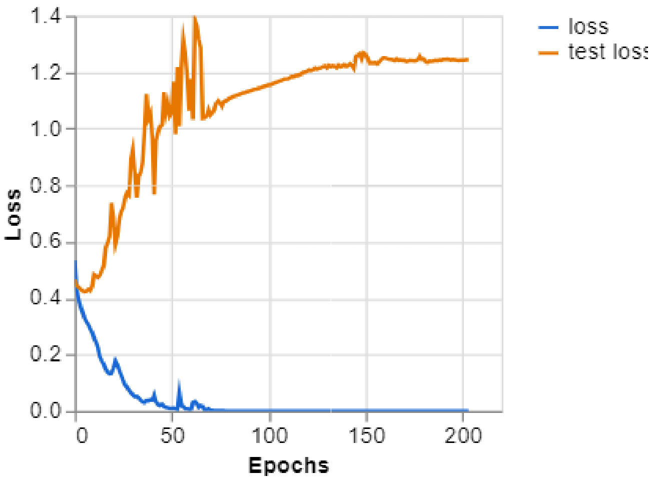


**Figure 6:** Loss function for pneumonia model. Testing in orange, and training in blue. Source: own results using Teachable Machine.

Accuracy per class

| CLASS | ACCURACY | # SAMPLES |
|---|---|---|
| Normal | 0.90 | 164 |
| Drusen | 0.87 | 160 |
| DME | 0.88 | 134 |
| CNV | 0.98 | 187 |

**Figure 7:** Accuracy per class. Legend: it is the percentage of right classification using a validation dataset. E.g., 0.9 means 90% of accurracy. Source: own results using Teachable Machine.

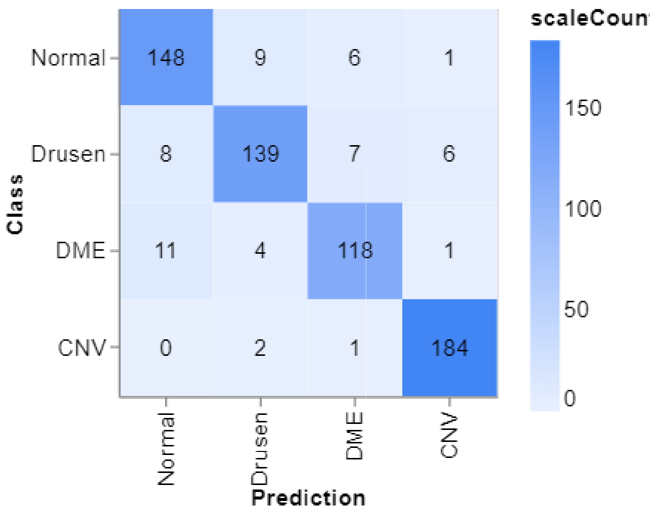


**Figure 8:** Confusion matrix to the retina model . Source: own results using Teachable Machine.

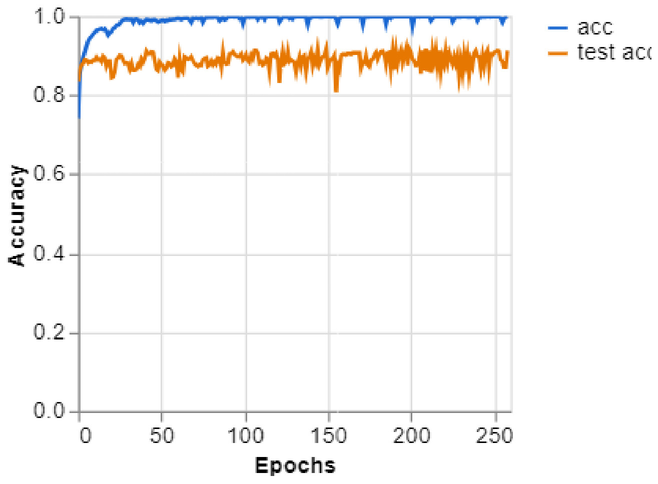


**Figure 9:** Accuracy graph during training for the retina model. **Legend**. final 99% for training (in blue) and 91% for testing (in orange). Source: own results using Teachable Machine.

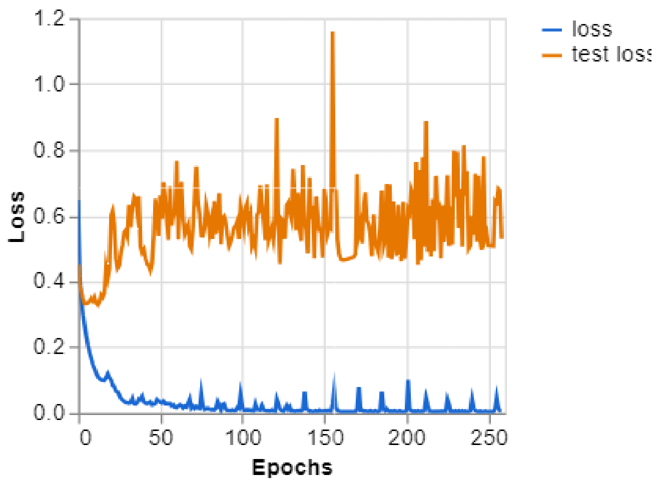


**Figure 10:** Loss function for retina model during training. Source: own results using Teachable Machine.

**URL:** *https://www.cell.com/cell/fulltext/S0092-8674(18)30154-5*

Laborde, G. (2021). *Learning Tensorflow.Js: Powerful Machine Learning in JavaScript*, O'Reilly Media, https://www.amazon.com.br/Learning-Tensorflow-Js-Powerful-Machine-JavaScript/dp/1492090794.

Pires, J. and Dias Braga, L. (2023). Snakeface: a transfer learning based app for snake classification, *Revista Brasileira de Computação Aplicada* **15**(3): 80–95. DOI: 10.5335/rbca.v15i3.15028.
**URL:** *https://seer.upf.br/index.php/rbca/article/view/15028*

Pires, J. G. (2023). Machine learning in medicine using javascript: building web apps using tensorflow.js for interpreting biomedical datasets, *medRxiv* .
**URL:** *https://www.medrxiv.org/content/early/2023/07/09/2023.06.21.23291717*
