## Supplementary material for "A conversational artificial intelligence based web application for medical conversations: a prototype for a chatbot": Robodoc: conversation demo repository

### Abstract

During the paper, the main manuscript, we have discussed the technical and scientific part of the chatbot we have built. We have built a conversational-AI for assisting patients/professionals on medical inquiries, triggered either by text or by image upload. Now, we present samples of conversations. Those samples were chosen to showcase some interesting behaviors that we believe deserve attention. We focus on the behaviors, not on the technical details. See the main manuscript for scientific and technical details on how the chatbot was built. We hope to give the reader a better feeling on how it is to interact with a conversational-AI designed to support on medical inquiries. The system can be triggered by either image upload or text entering, we are going to showcase both. For the image-triggered part, we are going to test each possible case, and showcase how the system behavior.

**Keywords**: chatbots; bioinformatics; conversational-AI; openAI; large language models; tensorflow.js; JavaScript; medicine; medical images; computer vision.

### 1 Samples of conversations with Robodoc: text-triggered model

On this section, we shall focus on the path that is triggered by text, by entering a message on the chatbot. This message should contain information from the patient, or the chatbot will ask for the information, or missing information.

#### 1.1 Configuration

The configuration is presented on Table 1.

#### 1.2 More information than needed

We have "randomly" picked one patient from our dataset, the complete dataset is here, with no diabetes.

> I have done a couple of tests, and I would like to know my chances of having diabetes. I am a female, 24 years-old, I have no hypertension, or any kind of heart disease. My BMI is 35.42, my HbA1c level is 4, and glucose level 100.

See that the current model does not use the gender, it was added to test whether the algorithm can ignore unnecessary information. We have actually shown that for this dataset the gender can make the difference Pires (2023a), something we may add to the model in the future.

> Robodoc: Based on the information you've provided, your chances of having diabetes are around 0.67% [which is correct, the patient has no diabetes]. It's always important to consult with a healthcare professional for a comprehensive assessment. If you have any other questions or need further assistance, feel free to ask!

Now, we just have one model. Therefore, no need to concern whether it calls the right model. But we can check the parameters entered, under the hood, those are the parameters it extracts from the text and passed to the diabetes model:

"{ "age": "24", "hypertension": 0, "heart_disease": 0, "bmi": "35.42", "HbA1c_level": "4", "blood_glucose_level": "100" }"

We have explored variations of this model on our previous paper Pires (2023b): we are using what is called the *6-feature model*. Those variations and more can be added to future versions of Robodoc. On this possible future scenario, the function calling technique will be in charge of smartly shifting between models according to the parameters passed to the bot.

We can even go on with the conversation, this is the interesting part. I have asked, on the same thread, tips to keep diabetes-free.

> Robodoc: "I'm happy to hear that you're proactive about your health! To help reduce the risk of diabetes, consider following some of these

**Table 1:** Configuration for the text-triggered option

| Stage | Model | obs. |
|---|---|---|
| parameter extracting and model picking | gpt-4-1106-preview | we had no choice, gpt-3.5-turbo-1106 was calling wrongly. |
| Final response | gpt-3.5-turbo-1106 | |

general tips: 1. Maintain a healthy weight and BMI. 2. Engage in regular physical activity and exercise. 3. Eat a balanced diet with plenty of fruits, vegetables, whole grains, and lean proteins. 4. Limit your intake of processed foods, sugars, and saturated fats. 5. Stay hydrated and choose water over sugary drinks. 6. Monitor your blood glucose levels if you have a history of high levels. 7. Avoid smoking and limit alcohol consumption. 8. Get enough sleep and manage stress effectively. 9. Have regular check-ups with your healthcare provider to monitor your health. Remember to always consult with a healthcare professional for personalized advice that considers your individual health needs and conditions. If you have specific questions or are looking for more details about managing your risk for diabetes, feel free to ask!"

#### 1.3 Missing information

We are going to use the same case, but we are going to remove one piece of information, namely, the blood glucose levels.

I have done a couple of testes, and I would like to know my chances of having diabetes. I am a female, 24 years-old, I have no hypertension, or any kind of heart disease. My BMI is 35.42, my HbA1c level is 4 [missing information]

Robodoc: To assess your chances of having diabetes using our diabetes_model tool, I need to know your blood glucose level. Could you please provide that information?

Me: Sure, glucose level 100.

Robodoc: thanks for all the information, give-me a second as I calculate for you.

Robodoc: Based on the information you provided, your chances of having diabetes are estimated to be approximately 0.67%. It's always important to discuss any concerns with a healthcare professional for personalized advice and further evaluation. Let me know if there's anything else I can assist you with!

Now, we have just one model, but we can build variations with different levels of confidence, as we have shown in Pires (2023b). Then, we allow the function calling technique to decide which model to call. This is something we need to test in the future. Based on what we have seen so far, we believe it can be done easily. It will require for sure prompt engineering, as we like to call the process of testing commands to those LLMs, and see their behavior, looking for a desired one.

### 2 Samples of conversations with Robodoc: image-triggered model

On this section, we are going to present the image-triggered path. This is automatically triggered when an image is uploaded, and all the process is triggered in cascade: from image description to final medical appointment . Different from the text-triggered path, this path has access to two medical image models, and three appointment sub functions (see the obs. column from Table 5 and Table 4). The medical image models are for X-ray and OCT images. They can diagnosis pneumonia (Table 5) and a couple of eye's conditions (Table 4).

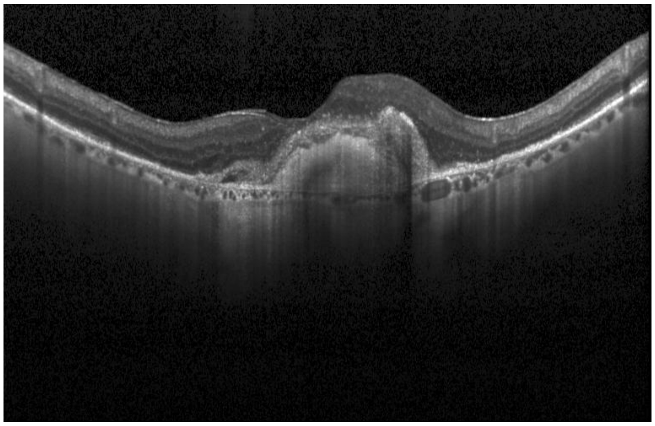

**Figure 1:** OCT with CNV. Source: OCT dataset, on the folderer 'test' (image: *CNV-5429703-1*).

#### 2.1 Configuration of the system

For deciding on the professional to make an appointment with, according to the diagnosis, we are using the suggestions from Kermany et al. (2018); see Table 4 and Table 5. The configurations for the conversation is on Table 2 and Table 3.

#### 2.2 Classes

The information on the tables was collected from Kermany et al. (2018).

The images used were all "randomly" selected: we just picked one on the test folders, they were not used during the training phase. We have used the same files from Kermany et al. (2018), including how they separated between training and testing.

#### 2.3 Optical Coherence Tomography (OCT)

##### 2.3.1 *Choroidal neovascularization (CNV)*

###### 2.3.1.1 *Information on the model under the hood behavior .*

- It called just the model *retinal_model* at the first stage, which is correct;
- CNV OCT by pretrained model by teachable machine with probability 100%, which is correct;
- On second stage, called just *urgent_referrals*, which is correct, it is an urgent appointment. Attached the observation for the medical doctor for the appointment "Presence of choroidal neovascularization (CNV) with a high probability." ;

No undesirable behavior on this case.

###### 2.3.1.2 *Chatbot content.*

**Table 2:** Configuration for the first stage: diagnosis from an image

| State | Algorithm | obs. |
|---|---|---|
| Image description | gpt-4-vision-preview | Creates an image description with possible diagnosis when possible |
| Choosing model | gpt-3.5-turbo-1106 | It will choose which model to call for making the diagnosis using computer vision |
| Making a final guess | gpt-3.5-turbo-1106 | It wil gather all information so far, and make a final diagnosis |

**Table 3:** Second stage: finding a professional accordingly.

| Stage | algorithm | obs. |
|---|---|---|
| Choosing the professional to call | gpt-3.5-turbo-1106 | Given a set of professional, decides which group to call. |
| Making an appoitment and informing the patient | gpt-3.5-turbo-1106 | We are using a stub: a dummy function. |

Robodoc: Welcome to our bot focused on medicine!. We're here to help!

Robodoc: See you uploaded an image.

Robodoc: Let me try one trick. I am analyzing the image for you.

Robodoc: This what I can see and say about this image.

Robodoc: The image appears to be an Optical Coherence Tomography (OCT) scan. OCT is a non-invasive imaging test that uses light waves to take cross-section pictures of your retina, the light-sensitive tissue lining the back of the eye. The scan can provide detailed images that can help with the diagnosis and monitoring of various eye conditions. However, I'm unable to diagnose medical conditions or provide clinical interpretations of medical images. For a proper diagnosis and medical advice, a qualified healthcare professional would need to evaluate the image in the context of the patient's history, symptoms, and other test results. If you have concerns about what this image might indicate, it is best to consult with an ophthalmologist or an eye care specialist.

: Robodoc: We are going to use a tool called retinal_model

Robodoc: The Optical Coherence Tomography (OCT) scan indicates the presence of choroidal neovascularization (CNV) with a high probability. It's important to consult with an ophthalmologist for a detailed evaluation and appropriate management.

Robodoc: You have an appointment with Dr. Sheldon Cooper, an ophthalmologist, who will support you from now on.

#### 2.3.2 *Diabetic Macular Edema (DME)*

##### 2.3.2.1 *Information on the model under the hood behavior .*

- It called just the model *retinal_model* at the first stage, which is correct;
- DME OCT by pretrained model by teachable machine with probability 100%, which is correct;
- On second stage, called just *routine_referrals*, which is wrong, it is an urgent appointment. Attached the observation for the medical doctor for the appointment "The patient has signs consistent with diabetic macular edema (DME). Please schedule a routine referral with an eye specialist for further evaluation and management.";

This case took several attempts, it tends to confuse with drusen. Also, it did not send to urgent. On the overall, the

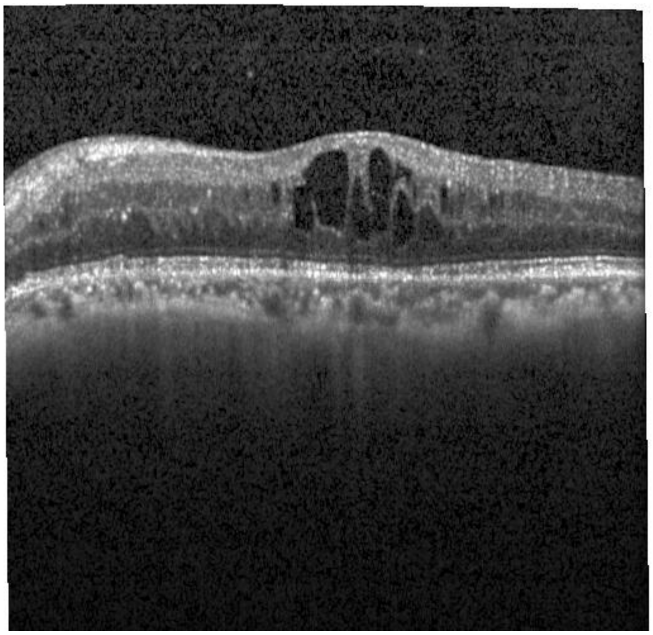

**Figure 2:** OCT with DME. Source: OCT dataset, on the folder 'test' (image: *DME-8615417-1*).

**Table 4:** Possible classifications for Optical Coherence Tomography (OCT) images

| Class | obs. |
|---|---|
| Choroidal Neovascularization | urgent referrals. |
| Diabetic Macular Edema | urgent referrals. |
| Drusen | routine referrals. |
| Normal | observation |

**Table 5:** Different classes of X-ray images with pneumonia.

| Class | obs. |
|---|---|
| normal | |
| bacterial | urgent referral for immediate antibiotic treatment |
| Virual | treated with supportive care |

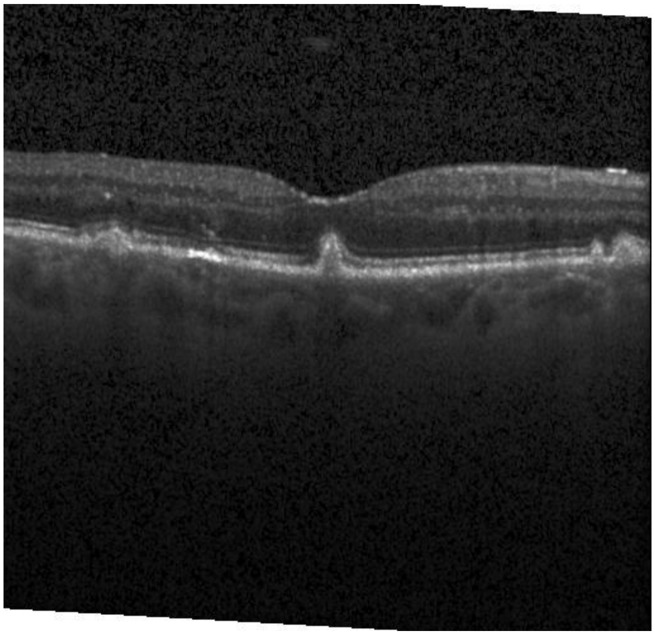

**Figure 3:** OCT with drusen. Source: OCT dataset, on the folder 'test' (image: *DRUSEN-9624303-1*).

behavior is okay.

##### 2.3.2.2 Chatbot content.

Robodoc: Welcome to our bot focused on medicine! . We're here to help!

Robodoc: See you uploaded an image.

Robodoc: Let me try one trick. I am analyzing the image for you.

Robodoc: This what I can see and say about this image.

Robodoc: The image appears to be an Optical Coherence Tomography (OCT) scan, which is a non-invasive imaging test often used by ophthalmologists to capture detailed images of the retina to aid in the diagnosis of retinal diseases. The black areas in the center of the image could indicate fluid accumulations or abnormalities in the retina's structure. Conditions such as macular degeneration, macular edema, macular hole, or central serous retinopathy often present with similar patterns on OCT scans. However, without a medical history, clinical context, or a professional medical assessment, it is not possible to provide a precise diagnosis. For a definitive diagnosis, this image should be evaluated by a qualified medical professional who would correlate these findings with clinical examination and possibly other diagnostic tests.

: Robodoc: We are going to use a tool called retinal_model

Robodoc: The image shows signs consistent with diabetic macular edema (DME). This condition involves the accumulation of fluid in the macula, which can lead to vision impairment. It is essential to consult with an ophthalmologist for further evaluation and management.

Robodoc: I have scheduled a routine referral for you to see an ophthalmologist regarding the signs consistent with diabetic macular edema (DME). You have an appointment with Dr. Sheldon Cooper, who will support you from now on.

#### 2.3.3 Drusen

##### 2.3.3.1 Information on the model under the hood behavior .

- It called just the model *retinal_model* at the first stage, which is correct;
- Drusen OCT by pretrained model by teachable machine with probability 100%, which is correct;

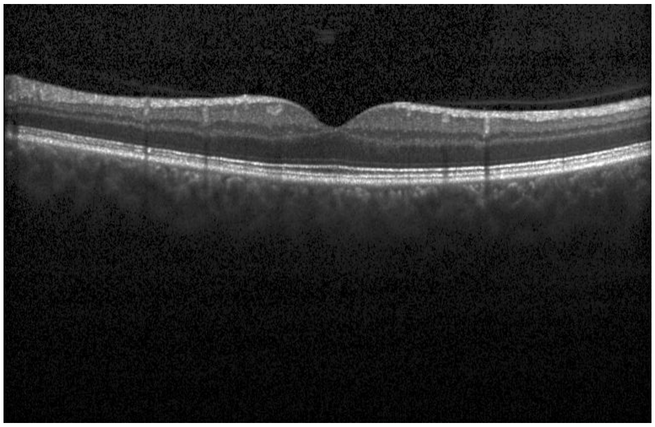

**Figure 4:** Normal OCT. Source: OCT dataset, on the folder 'test' (image: *NORMAL-626127-1*).

- On second stage, called just *routine_referrals*, which is correct. Attached the observation for the medical doctor for the appointment "Presence of drusen in the retina with high confidence. ";

Therefore, no undesired behavior was observed on those simulations.

##### 2.3.3.2 Chatbot content.

Robodoc: Welcome to our bot focused on medicine!. We're here to help!

Robodoc: See you uploaded an image.

Robodoc: Let me try one trick. I am analyzing the image for you.

Robodoc: This what I can see and say about this image.

Robodoc: The image appears to be an Optical Coherence Tomography (OCT) scan of a human retina. OCT is a non-invasive imaging test that uses light waves to take cross-section pictures of the retina, which is the light-sensitive tissue lining the back of the eye. The scan seems to show the different layers of the retina with good definition. Normally, an OCT scan is used to help diagnose conditions such as macular degeneration, glaucoma, retinal detachment, macular holes, and more. As for guessing a medical diagnosis, it would be inappropriate and potentially inaccurate for me to attempt to diagnose a medical condition based on this image without the relevant medical qualifications and the full clinical context. It is always best to consult a healthcare professional for an accurate diagnosis and advice.

Robodoc: We are going to use a tool called retinal_model

Robodoc: The model has identified the presence of drusen in the retina with high confidence. Drusen are small yellow deposits under the retina and are often associated with age-related macular degeneration. It's important to consult an eye care professional for further evaluation and management.

Robodoc: I have scheduled an appointment with Dr. Sheldon Cooper, an ophthalmologist, who will further evaluate the presence of drusen in your retina and provide necessary management.

#### 2.3.4 Normal

#### 2.3.5 Information on the model under the hood behavior

- It called just the model *retinal_model* at the first stage, which is correct;
- Normal OCT by pretrained model by teachable machine with probability 100%, which is correct;
- On second stage, called just *observation_referrals*, which is correct. Attached the observation for the medical doctor for the appointment "The retinal image suggests

a normal retina with a high probability of 99.94%.";

Therefore, no undesired behavior was observed on those simulations.

##### 2.3.6 *Chatbot content*

Robodoc: Welcome to our bot focused on medicine! . We're here to help!

Robodoc: See you uploaded an image.

Robodoc: Let me try one trick. I am analyzing the image for you.

Robodoc: This what I can see and say about this image.

Robodoc: The image appears to be an optical coherence tomography (OCT) scan, which is a non-invasive imaging test often used in ophthalmology to capture detailed images of the retina. This is the part of the eye that is responsible for capturing images from the visual field. The scan shows a cross-sectional view of the retina, which includes various layers that should be intact and well-differentiated in a healthy eye. In this particular image, it's possible to notice a central dipping or depression in the retina's profile. Such a feature could be consistent with a condition known as macular hole or macular edema, but other retinal conditions could also be a possibility. It's important to remember that interpreting medical images like OCT scans requires professional expertise in ophthalmology, and a diagnosis would typically involve a comprehensive examination including a review of the patient's medical history, symptoms, and additional testing as needed. Therefore, while some features can be suggestive of certain conditions, a definitive diagnosis should always be provided by a qualified medical professional.

Robodoc: We are going to use a tool called $retinal_model$

Robodoc: The analysis of the retinal image suggests that the retina appears to be normal, with a high probability of 99.94%. However, it's important to consult with an ophthalmologist for a comprehensive evaluation and interpretation of the findings.

Robodoc: You have been scheduled for an appointment with Dr. Sheldon Cooper, who will provide professional support and further evaluate the retinal image findings.

### 2.4 Pediatric pneumonia

On this section, we shall test the model for images with X-ray from the lungs. We are going to consider each possible case on the current prototype.

##### 2.4.1 *Bacterial pneumonia*

Bacterial pneumonia is a type of lung infection caused by bacteria. It can affect one or both lungs and cause symptoms such as cough, fever, chest pain, and difficulty breathing. Bacterial pneumonia can be serious and sometimes life-threatening, especially in people with weakened immune systems, chronic diseases, or older age. Bacterial pneumonia is usually treated with antibiotics, but some bacteria are resistant to certain drugs. Therefore, it is important to seek medical attention as soon as possible if you suspect you have bacterial pneumonia. We following the suggestions from Kermany et al. (2018), this is an urgent case, and should trigger a urgent appointment request (Table 5).

##### 2.4.2 *Information on the model under the hood behavior*

- It called just the model *pneumonia_model* at the first stage, which is correct;
- Bacterial pneumonia by pretrained model by teachable machine with probability 99%, which is correct;
- On second stage, called just *urgent_referrals_pneumonia* , which is correct. Attached the observation for the

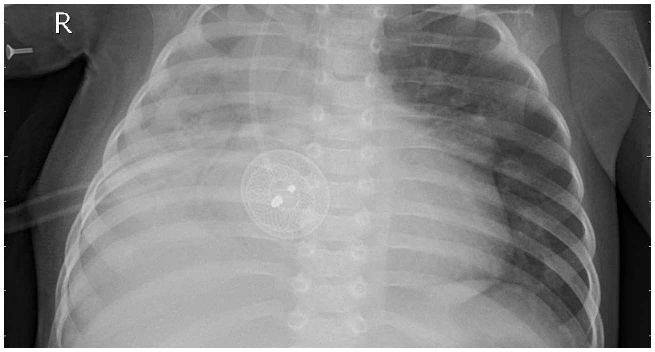


**Figure 5:** Bacterial Pneumonia. Source: X-ray dataset, on the folder 'test' pneumonia (image: *person85_bacteria_422*).

medical doctor to the appointment "High probability of bacterial pneumonia based on chest X-ray analysis";

Therefore, no undesired behavior was observed on those simulations.

##### 2.4.3 *Chatbot content*

Robodoc: Welcome to our bot focused on medicine!. We're here to help!

Robodoc: See you uploaded an image [image uploading triggers a bot response].

Robodoc: Let me try one trick. I am analyzing the image for you [this is GPT 4 making an image description].

Robodoc: This what I can see and say about this image [GPT 4 description].

Robodoc: The image you provided appears to be a chest X-ray. However, I'm not able to offer medical diagnoses or healthcare advice. For interpretation of radiological images and medical diagnoses, it's important to consult with a trained healthcare professional who can give an accurate assessment based on their expertise and the clinical context of the patient. What I can note from the X-ray, assuming it is for illustrative purposes:

1. There are what appear to be coin-like objects overlying the area of the stomach. This might suggest an ingestion or aspiration of foreign bodies, which is a medical concern that typically requires evaluation by a healthcare provider.

2. The lung fields, heart silhouette, bony structures, and diaphragm are visible, which are usually assessed in chest X-rays for signs of abnormalities such as pneumonia, tuberculosis, heart enlargement, fractures, or other conditions. If this chest X-ray relates to a real-life medical case, it is crucial for the individual in question to seek professional medical evaluation.

Robodoc: We are going to use a tool called pneumonia_model.

Robodoc: Based on the analysis of the chest X-ray, it appears that there is a high probability of bacterial pneumonia present [this is after function calling, under the hood, it called the pneumonia model we trained, and added the result to its final response, 99% of bacterial pneumonia]. It's important for the individual to seek medical attention and further evaluation from a healthcare professional.

Robodoc: You have an appointment with Dr. Sheldon Cooper, a pneumonia specialist, who will support you from now on. [this is the second stage, and final, it uses all the information so far to make an appointment with a professional]

This conversation was perfect, no undesired behavior seen. We did not have to try several times, this is the first attempt.

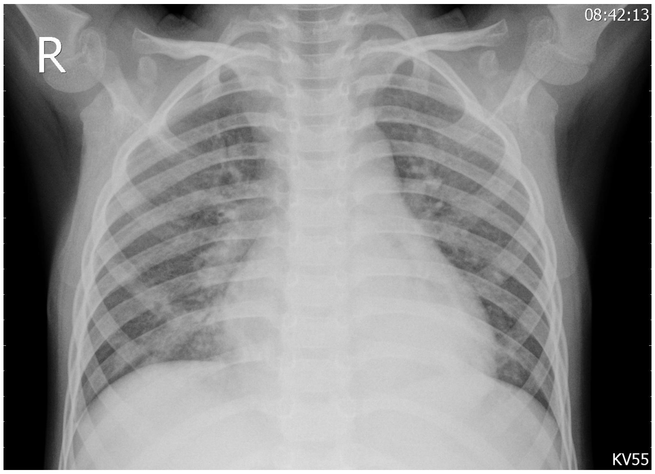


**Figure 6:** Viral pneumonia. Source: X-ray dataset, on the folder 'test' pneumonia (image: *person78_virus_140*).

##### *2.4.4 Viral pneumonia*

Viral pneumonia is a type of lung infection caused by viruses. It can affect one or both lungs and cause symptoms such as cough, fever, shortness of breath, and chest pain. Viral pneumonia can be mild or severe, depending on the type of virus and the person's health status. Some common causes of viral pneumonia are influenza, respiratory syncytial virus (RSV), and coronavirus. Viral pneumonia is diagnosed by a physical exam, chest X-ray, and sometimes a blood test or a nasal swab. Treatment for viral pneumonia depends on the cause and the severity of the illness. Some viral infections may resolve on their own, while others may require antiviral medications or hospitalization. Prevention of viral pneumonia includes getting vaccinated against influenza and other viruses, washing hands frequently, avoiding contact with sick people, and wearing a mask when needed. We following the suggestions from Kermany et al. (2018), this is not an urgent case, and should not trigger a urgent appointment request (Table 5).

##### *2.4.5 Information on the model under the hood behavior*

- It called just the model *pneumonia_model* at the first stage, which is correct;
- Viral pneumonia by pretrained model by teachable machine with probability 100%, which is correct;
- On second stage, called just *urgent_referrals_pneumonia* , which is correct; nonetheless, viral pneumonia is not considered urgent (see Table 5). Attached the observation for the medical doctor to the appointment "Chest X-ray shows evidence of viral infection. Urgent referral for evaluation and treatment.";

Therefore, just one undesired behavior was observed on those simulations. However, it does not seem to be serious. We believe that testing different prompts may improve the function calling. We have seen similar behaviors as we created the assistant, and by trying out different prompts, we were able to get what we wanted. This is a prompt engineer issue.

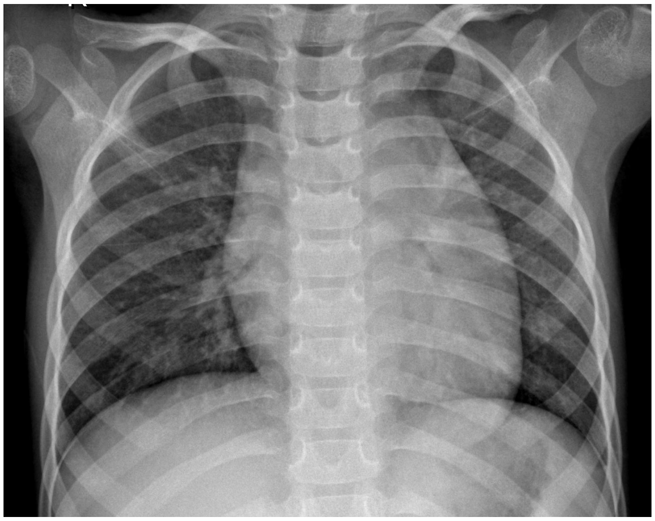

**Figure 7:** Normal X-rays. Source: X-ray dataset, on the folder 'test' pneumonia (image: *NORMAL2-IM-0035-0001*).

##### *2.4.6 Chatbot content*

Robodoc: Welcome to our bot focused on medicine!. We're here to help!

Robodoc: See you uploaded an image.Robodoc: Let me try one trick. I am analyzing the image for you.

Robodoc: This what I can see and say about this image.

Robodoc: This image appears to be a chest X-ray. Unfortunately, as an AI, I'm not capable of providing medical diagnoses. For an accurate diagnosis and analysis, this image should be reviewed by a qualified medical professional, such as a radiologist, who is trained to interpret such images and provide accurate clinical correlations and diagnoses based on the full clinical context of the person in question. If you have concerns about what this X-ray might indicate, it is important to consult a healthcare provider.

Robodoc: We are going to use a tool called pneumonia_model

Robodoc: The chest X-ray shows evidence of a viral infection. It's important to consult a healthcare professional for further evaluation and appropriate management.

Robodoc: I have made an urgent referral for you to see Dr. Sheldon Cooper, a pneumonia specialist, to evaluate and treat the viral infection indicated by your chest X-ray. Please make sure to follow up with the appointment as soon as possible.

##### *2.4.7 Normal X-ray*

##### *2.4.8 Information on the model under the hood behavior*

- It called just the model *pneumonia_model* at the first stage, which is correct;
- Normal by pretrained model by teachable machine with probability 100%, which is correct;
- On second stage, called just *observation_referrals* , which is correct. Attached the observation for the medical doctor to the appointment "The chest X-ray appears normal .";

On this case, we had to try several times to get one right. It tends to classify as pneumonia, either viral or bacterial. This mistake is better than the opposite: classifying as normal a patient with pneumonia. This behavior is curious since the confusion matrix and error per class did not show that. One possibility is that the images from the testing dataset are from a different X-ray machine. We are using the same separation from Kermany et al. (2018).

Robodoc: Welcome to our bot focused on medicine! . We're here to help!

Robodoc: See you uploaded an image.

Robodoc: Let me try one trick. I am analyzing the image for you.Robodoc: This what I can see and say about this image.

Robodoc: This is an image of a chest X-ray. The image is a radiograph that typically allows medical professionals to examine the condition of the lungs, heart, and bones of the spine and chest. As an AI, I cannot make an accurate medical diagnosis, and it would not be appropriate to guess a medical diagnosis based on an image. Medical diagnoses should be made by qualified healthcare professionals who can interpret medical images in the context of a patient's history, symptoms, physical examination, and other diagnostic tests. If you have concerns about what this image might indicate, I strongly advise consulting a medical professional for an accurate and reliable interpretation.:

Robodoc: We are going to use a tool called pneumonia_model

Robodoc: The chest X-ray appears normal, indicating that there are no signs of pneumonia in the lungs. If you have any concerns about your health, I recommend consulting a healthcare professional for further evaluation and guidance.

Robodoc: You have an appointment with Dr. Sheldon Cooper, an specialist, who will support you from now on. If you have any further concerns about your health, please feel free to discuss them with Dr. Cooper during your appointment.
